# Point-of-Care MRI with Artificial Intelligence to Measure Midline Shift in Acute Stroke Follow-Up

**DOI:** 10.1101/2022.01.22.22269697

**Authors:** Prantik Kundu, Seyed Sadegh Mohseni Salehi, Bradley A. Cahn, Mercy H. Mazurek, Matthew M. Yuen, E. Brian Welch, Barbara S. Gordon-Kundu, Jo Schlemper, Gordon Sze, W. Taylor Kimberly, Jonathan M. Rothberg, Michal Sofka, Kevin N. Sheth

## Abstract

**Background and Purpose:** In stroke, timely treatment is vital for preserving neurologic function. However, decision-making in neurocritical care is hindered by limited accessibility of neuroimaging and radiological interpretation. We evaluated an artificial intelligence (AI) system for use in conjunction with bedside portable point-of-care (POC)-MRI to automatically measure midline shift (MLS), a quantitative biomarker of stroke severity.

**Materials and Methods:** POC-MRI (0.064 T) was acquired in a patient cohort (n=94) in the Neurosciences Intensive Care Unit (NICU) of an academic medical center in the follow-up window during treatment for ischemic stroke (IS) and hemorrhagic stroke (HS). A deep-learning architecture was applied to produce AI estimates of midline shift (MLS-AI). Neuroradiologist annotations for MLS were compared to MLS-AI using non-inferiority testing. Regression analysis was used to evaluate associations between MLS-AI and stroke severity (NIHSS) and functional disability (mRS) at imaging time and discharge, and the predictive value of MLS-AI versus clinical outcome was evaluated.

**Results:** MLS-AI was non-inferior to neuroradiologist estimates of MLS (p<1e-5). MLS-AI measurements were associated with stroke severity (NIHSS) near the time of imaging in all patients (p<0.005) and within the IS subgroup (p=0.005). In multivariate analysis, larger MLS-AI at the time of imaging was associated with significantly worse outcome at the time of discharge in all patients and in the IS subgroup (p<0.05). POC-MRI with MLS-AI >1.5 mm was positively predictive of poor discharge outcome in all patients (PPV=70%) and specifically in patients with IS (PPV=77%).

**Conclusion:** The integration of portable POC-MRI and AI provides automatic MLS measurements that were not inferior to time-consuming, manual measurements from expert neuroradiologists, potentially reducing neuroradiological burden for follow-up imaging in acute stroke.

## 1. Introduction

Stroke continues to be a leading cause of death and disability in people of all ages worldwide (GBD 2015 Neurological Disorders Collaborator Group, 2017; Dewan et al., 2018). The monitoring of imaging biomarkers, such as midline shift (MLS), that signify neurological damage after stroke is one of the most prominent challenges in neurointensive care. Conventional imaging techniques used to monitor these biomarkers, such as computed tomography (CT) and magnetic resonance imaging (MRI), typically require transportation of patients from the intensive care unit to a different location within the hospital, which greatly increases the risk of complications (Jia et al., 2016; Parmentier-Decrucq et al., 2013; Smith et al., 1990).

Point-of-care (POC) versions of conventional imaging modalities, such as transcranial doppler (TCD) ultrasound (Blanco and Abdo-Cuza, 2018; Lau and Arntfield, 2017), computed tomography (POC-CT) (LaRovere et al., 2012; Peace et al., 2010), and low-field POC magnetic resonance imaging (POC-MRI) (Cooley et al., 2021; Sheth et al., 2020; Turpin et al., 2020), are emerging as potential solutions to increase the availability of imaging at the patient bedside, potentially revolutionizing neurocritical care workflows 1/22/22 4:18:00 PM. However, each of these modalities has its own limitations. TCD is operator-dependent and limited by the size and location of the acoustic windows – regions of where the skull is thin enough for ultrasound to penetrate (Naqvi et al., 2013). POC-CT has inherently low soft-tissue contrast and exposes patients to ionizing radiation (Rumboldt et al., 2009). While the image resolution and number of sequences available are currently limited in POC-MRI relative to conventional, high-field MRI, POC-MRI overcomes the limitations of TCD and POC-CT by offering whole-brain images with excellent soft-tissue contrast that are acquired without ionizing radiation and are not operator-dependent. A preliminary study of neurointensive care patients with neurologic symptoms owing to severe COVID-19 and stroke pathology demonstrated the sensitivity of POC-MRI to neuropathophysiology (Sheth et al., 2020).

One of the benefits of bringing imaging to the patient’s bedside in a neurocritical care unit is the speed with which images can be acquired; however, this benefit may be negated if the clinicians must wait for an official read or analysis of the imaging biomarkers from another department. For example, MLS measurements typically require manual definition of anatomical landmarks or evaluation of images in separate software packages (Liao et al., 2018). Artificial intelligence (AI) provides a mean for capturing expert knowledge in an automated image assessment algorithm that has been trained using input from experts and can be incorporated directly into the image workflow.

AI serves to augment decision making by automating the interpretation of data, thereby complementing or supplementing human evaluation (Hainc et al., 2017). Deep learning (DL)—a sub-type of AI— has been used in medical imaging for pathology detection and classification, as well as image filtering (Serag et al., 2019; Taghanaki et al., 2021; Vieira et al., 2017). In supervised DL, artificial neural network models are trained through minimizing the error of predicting ‘ground truth’ features of interest in training data (LeCun et al., 2015). The combination of POC-MRI and automated biomarker assessment holds great potential for improving the workflow in the neurocritical care setting.

The primary aim of our study was to compare the performance of a supervised DL algorithm trained to automatically measure MLS to that of manual assessment by expert neuroradiologists from POC-MRI data acquired in a cohort of stroke patients in neurocritical care. Additionally, the relationship between the automated MLS measures and clinical outcomes for the patients was assessed.

## 2. Materials and Methods

### 2.1. Patients

Our study was conducted under an institutional review board (IRB) protocol approved by the Yale Human Research Protection Program, and written informed consent was obtained from all participants or their legally authorized representatives prior to any research activities.

Between July 2018 to March 2020, all patients who were admitted for stroke to the Neurosciences Intensive Care Unit (NICU) at the Yale New Haven Hospital and had visible brain pathology on conventional neuroimaging (CT or high-field MRI) were screened for the study. Inclusion criteria included age ≥ 18 years, admission to the NICU for stroke, and visible brain pathology on standard of care imaging. Exclusion criteria included code status (i.e., patients that were not clinically stable), isolation requirements (e.g., due to MRSA, c. diff, or e. coli), or the presence of at least one of the following MRI contraindications: cardiac pacemakers or defibrillators, intravenous medication pumps, insulin pumps, deep brain stimulators, vagus nerve stimulators, cochlear implants, pregnancy, and cardiorespiratory instability.

Patient age, primary diagnosis, stroke severity, and discharge outcomes were recorded, if available. Primary diagnoses were either ischemic stroke (IS) or hemorrhagic stroke (HS), and the HS group was subdivided into intraparenchymal hemorrhage (IPH) and subarachnoid hemorrhage (SAH). Stroke severity was measured by the NIH stroke scale (NIHSS, score: 0–42) and recorded at the time of POC-MRI imaging. Discharge outcomes were recorded at the time of discharge and at 90-day follow-up, if available, using the modified Rankin Scale (mRS; 0–6), which captures the degree of disability or dependence in daily activities of stroke patients or other causes of neurological disability, where an mRS score of 6 indicates expiry (Ostwaldt et al., 2018; Quinn et al., 2009; Ropper, 1986; Sulter et al., 1999).

### 2.2. Portable POC-MRI imaging

Patient imaging was performed during the follow-up period after initial treatment for acute stroke. Images were acquired at bedside in the Neuro ICU using an FDA-cleared, ultra-low magnetic field (64 mT) portable POC-MRI system (Swoop(tm), Mk 1.2 RC6.3–7.2 software; Hyperfine, Inc., Guilford, CT, USA) with an 8-channel head coil and a biplanar 3-axis gradient system with peak amplitudes of 26 mT/m (Z-axis) and 25 mT/m (X- and Y-axis). Patients were positioned in the head coil inside the imaging area of the portable POC-MRI while in standard hospital beds (Figure 1a). Ongoing standard of care treatment (i.e., ventilation, intravenous infusions, and telemetry) continued during the imaging exam, and radiofrequency interference cancellation was enabled on the POC-MRI system (Rearick et al., 2017). T_2_-weighted POC (T2W_POC_) images were acquired: repetition time (TR) = 4000 ms; echo time (TE) = 228 ms; inversion time (TI) = 1400 ms; 1.5 × 1.5 × 5 mm^3^ resolution; 36 slices; and an approximately 5 min scan duration.

**Figure 1:**
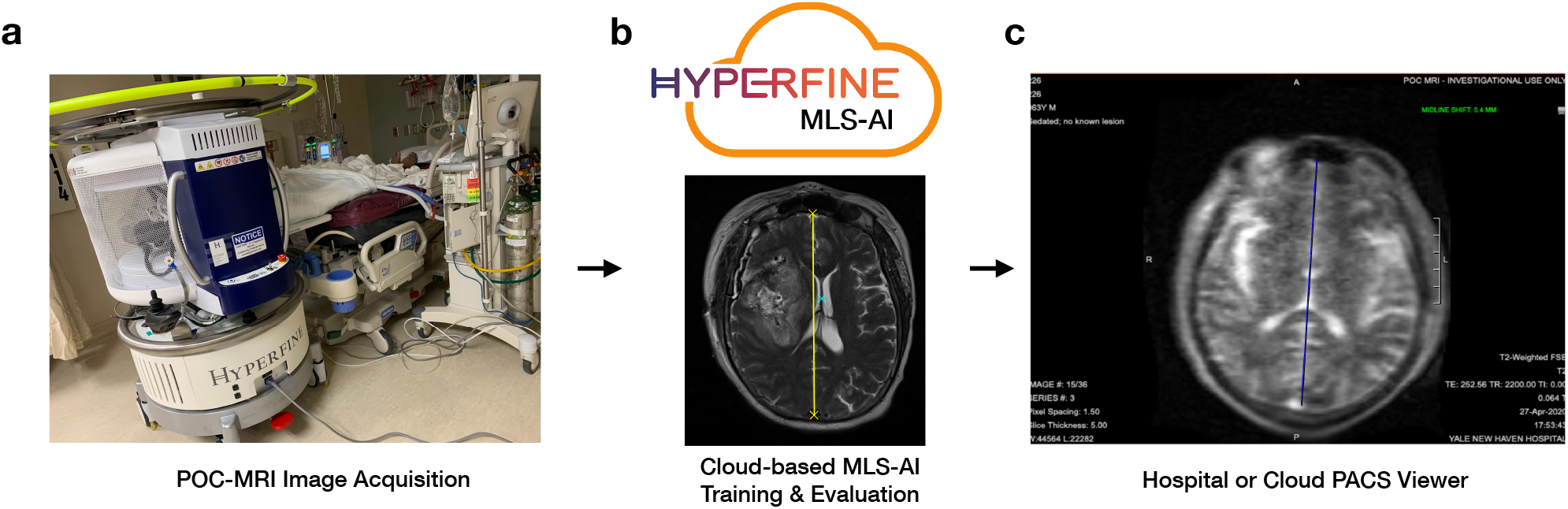
(a) Bedside imaging of a patient during treatment using the POC 64mT MRI system (Hyperfine, Inc. Swoop, Mk 1.2 RC6.3–7.2 software). (b) Example annotation of brain midline on an example 1.5 T dataset (collected separately) using anterior and posterior falx cerebri as endpoints (yellow) and septum pellucidum as the shift point (cyan cross). (c) After processing, POC-MRI images annotated by MLS-AI were viewable on the vendor-provided cloud PACS.

All image data were deidentified as part of the IRB-approved protocol, and no personally identifiable information (PII) was accessible in this study. Imaging data were uploaded to a cloud picture archiving and communication system (PACS) for further analysis

### 2.3. Additional data sets

#### 2.3.1. Training data sets

Two additional data sets were used to train the MLS-AI models. First, high-field T2W (T2W_HF_) images publicly available from the Human Connectome Project (n=528) were adapted to match the T2W_POC_ image resolution and noise content (Van Essen et al., 2013). The T2W_HF_ images were acquired at 3.0 T with the following parameters: TR = 3200 ms; TE = 565 ms; 0.7 × 0.7 × 0.7 mm^3^ resolution, and a scan duration of 8 min 24 s. Second, low-field T2W (T2W_LF_) images from the Hyperfine image archival system (n=86) were used. These de-identified images were acquired using the POC-MRI system (Swoop, Mk 1.2 RC6.3–7.2 software; Hyperfine, Inc., Guilford, CT, USA) at a variety of sites and represent a variety of unknown pathologies.

#### 2.3.2. Evaluation data set

For use in model evaluation only, low-field T2W images from healthy controls (n=10; T2W_LFHC_) were extracted from the Hyperfine image archival system. These images were acquired under a protocol approved by the New England IRB, and written informed consent was obtained from each participant prior to imaging. Participants were adults, aged 18 years old or older, with a body habitus compatible for scanning inside the POC-MRI. Exclusion criteria included contraindications for MRI and pregnancy. Imaging was performed at Hyperfine, Inc. (Guilford, CT) with a POC-MRI system (Swoop, Mk 1.2 RC6.3–7.2 software; Hyperfine, Inc.; Guilford, CT, USA).

### 2.4. Manual annotations and MLS estimation

Three independent neuroradiologists (3-5 years of experience each) annotated each image volume included in this study (T2W_POC_, T2W_LF_, T2W_HF_, and T2W_LFHC_) using ITK SNAP (Yushkevich et al., 2016) software provided by study investigators. Each annotator selected the voxel corresponding to each of the three neuroanatomical landmarks used to estimate MLS (Figure 1b): the anterior and posterior falx cerebri and the septum pellucidum at the location of the largest shift from the midline at the slice level. A probability map was generated by convolving the map of the annotation points with a 3-D Gaussian kernel (σ=6 mm). The concatenation of the 3-D probability maps into a 4-D dataset from a single annotator comprised an annotation data set, resulting in three annotation data sets for each image volume.

MLS measurements were obtained by drawing a line from the anterior and posterior attachments of the falx cerebri, drawing a second, perpendicular line to the septum pellucidum at the point of maximal deviation, and measuring in millimeters the length of the second line.

### 2.5. Automated MLS estimation

MLS-AI estimates were derived from each patient data set (i.e., T2W_POC_) with a commercially available AI system (BrainInsight, Hyperfine Research Inc, Guilford CT) using an end-to-end fully convolutional neural network (CNN) based on the 3-D ResUNet architecture (Supplemental Figure 2) (Ronneberger et al., 2015).

**Figure 2:**
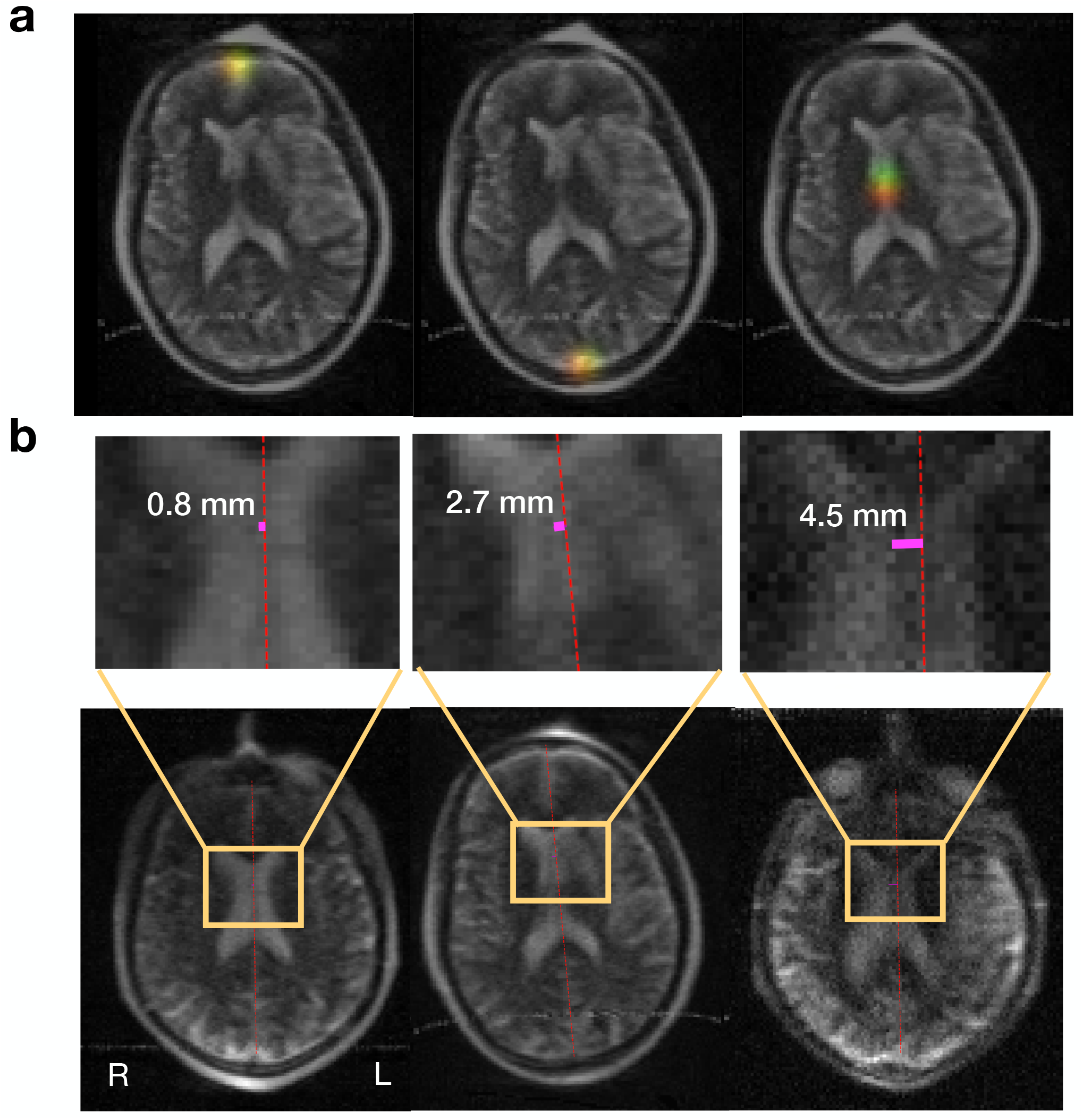
(a) Visualization of T2-weighted POC-MRI with probability density functions (i.e., heatmaps) for the target anatomical points of the corresponding landmarks. Red distributions correspond to ground truth locations (based on human annotation), green distributions are artificial intelligence-based MLS (MLS-AI) estimates of the target points, and yellow represents the overlap of the ground truth and estimated distributions. (b) MLS-AI estimates of stroke brain MLS graphically are overlaid on anatomical volumes in POC-MRI T2-weighted images.

To produce models representative of the image quality of the acquired stroke data set, a two-fold cross-validation experiment, involving model training and evaluation, was performed using the T2W_POC_, T2W_LF_, and T2W_HF_ image volumes and their corresponding annotations. Each independent fold was composed of half of the T2W_POC_ data (randomly split) and all the T2W_LF_ and T2W_HF_ data sets. The images and annotations in each fold were augmented to further increase the variation present in the data set through random geometric distortions (see Supplemental Information) and then used to train an MLS-AI model. The data sets within each fold were further subdivided, with 80% used for model training and 20% for model validation. Training was conducted in steps using batches of training data, with each training step followed by a validation step. The validation step was used to determine if a model updated with a batch of training data was more predictive of ground truth in the independent validation data, in which case the model was updated, otherwise not. (A detailed description of the MLS-AI model is provided in Supplemental Information.) The resulting model was then used to evaluate the T2W_POC_ images in the opposing fold.

In evaluation, MLS-AI model inputs were the 3D T2W POC-MRI volumes (i.e., T2W_POC_) and outputs were probability maps for the three anatomical landmarks used clinically to determine MLS (Figure 2a). The final MLS-AI estimate was computed from the voxels of peak probability for MLS landmarks, as the geometric displacement of the septum pellucidum to the brain mid-line drawn between the anterior and posterior falx (Figure 2b). No image was used in both training and evaluation of the same model.

To establish a background distribution of MLS-AI on POC-MRI from healthy controls, MLS-AI was also calculated using the T2W_LFHC_ data as input and the average of the models generated above.

### 2.6. Training accuracy of MLS-AI

To estimate the accuracy of the trained MLS-AI model against MLS estimates from human annotators for each image volume, the absolute difference of the MLS-AI estimate from the average human MLS estimate of all three annotators was computed. To accommodate variations in brain size, the difference was then normalized to the length of the brain midline. This measure of accuracy was calculated during the validation phase of training and for the final evaluation of the POC data.

### 2.7. Intra-rater and inter-method annotation discrepancy

Intra-rater landmark location discrepancy and the discrepancy between landmark locations defined by humans and MLS-AI were calculated as the mean absolute error (MAE) measured in millimeters between the landmark annotations and ground truth locations: MAE_Human_ and MAE_MLS-AI_, respectively. (See the Supplemental Information for additional details.) Since comparison to “true” values of radiologic measures is intractable, ground truth locations for each of the three landmarks were established on a relative basis. For the human annotations, ground truth was defined as the mean of the landmark locations from the other two annotators. For MLS-AI annotations (i.e., the locations of the peak probability), ground truth was defined as the mean of the landmark locations from three human annotators.

### 2.8. Qualitative threshold for MLS

To determine a threshold for dichotomizing MLS-AI to produce a qualitative marker for the presence or absence of MLS, values of MLS-AI at the minimum dimension of image resolution were evaluated for predictive value versus clinical outcome. A logistic regression was performed to evaluate associations between qualitatively worse outcome (mRS>3) and quantitative measurements of MLS-AI (Supplementary Figure 2). Values of 1.0 mm, 1.5 mm (the minimum image resolution), and 2.0 mm were evaluated for predictive value.

### 2.9. Statistical analyses

A paired-t-test was performed to compare MAE_MLS-AI_ and MAE_Human_. A two-tailed Student’s t-test was used to compare between the means of distributions of MLS-AI across study groups. The Mann-Whitney U-test was performed to compare rank-sum differences in MLS-AI across study groups. A hypothesis test for non-inferiority of MLS-AI to human annotators was conducted (Walker and Nowacki, 2011). The test compared the MLS-AI model discrepancy (MAE_MLS-AI_) to the average annotator discrepancy (MAE_Human_), as a fraction of average clinical annotator discrepancy, upper bounded by a clinically acceptable relative error, δ=0.2. Non-inferiority was established by showing that MAE_MLS-AI_ was significantly less than (1 + δ) MAE_Human_ at the α=0.05 significance level (see Supplemental Information for further details).

The relationship between MLS-AI estimates and stroke severity (NIHSS score) and disability at discharge (mRS score) was evaluated using linear regressions for all patients and in IS and HS, separately. A multivariate regression was performed controlling for patient age, as well. mRS was modeled as a dependent variable of MLS-AI. Additionally, we evaluated the qualitative effect of MLS-AI on patient outcome at discharge for IS and HS (binary logistic regression; mRS>3). In a sub-sample of patients with available follow-up clinical data, the relationship between MLS-AI mRS scores at 90 days post-discharge was examined with linear regression.

Ordinary and logistic regression analyses were conducted using *statsmodels* in Python (Seabold and Perktold, 2010). Leave-one-out cross-validation was conducted to determine the 95% percentile CIs of the estimated effect size. Regression analysis produced the regression coefficient, posterior probability (*p*), and 95% confidence intervals (two-tailed) using leave-one-out cross-validation. P-values less than 0.05 were considered statistically significant

## 3. Results

### 3.1. Patient demographics

A total of 94 patients were scanned with POC-MRI. The average patient was 62 years old and exhibited moderate stroke severity at the time of imaging (mean NIHSS=5) and moderate disability at the time of discharge (mean mRS=3; able to walk independently) Patients with data on age, diagnosis, severity and disability data (n=71) were grouped by primary diagnosis and stroke category: IS (n=38) and HS (n=33 total) with IPH (n=18) and SAH (n=15). Patient age, diagnosis, and stroke severity and disability scores are summarized in Table 1. No adverse events related to POC-MRI were reported.

**Table 1:** Patient demographics.

|  | Patient age<br>(years) |  | Stroke severity<br>(NIHSS; 1–42) |  | Disability at<br>discharge<br>(mRS; 1–6) |  |
| --- | --- | --- | --- | --- | --- | --- |
| | n | mean ± SD | n | mean ± SD | n | mean ± SD |
| All patients | 94 | 62 ± 16 | 92 | 6 ± 8 | 93 | 3 ± 2 |
| IS | 38 | 65 ± 18 | 46 | 8 ± 9 | 45 | 3 ± 2 |
| HS | 33 | 61 ± 13 | 38 | 4 ± 6 | 48 | 3 ± 2 |
| IPH | 18 | 62 ± 14 | 30 | 4 ± 5 | 31 | 3 ± 2 |
| SAH | 15 | 60 ± 13 | 8 | 2 ± 6 | 17 | 2 ± 2 |

HS: hemorrhagic stroke, IS: ischemic stroke, IPH: intraparenchymal hemorrhage, SAH: subarachnoid hemorrhage, NIHSS: NIH stroke severity score (1–42), mRS: modified ranking score (i.e., six-point disability scale, where 6 = expiry). Data are expressed as mean ± standard deviation (SD). Note that age, mRS, and NIHSS were not available for all patients imaged.

### 3.2. Training accuracy of MLS-AI

For each fold, the included image data sets were split 80%/20% for training/validation, respectively: T2W_POC_ (38/9), T2W_LF_ (66/20), and T2W_HF_ adapted to POC-MRI resolution and noise content (470/58). The accuracy of the MLS-AI estimates was 20.7 ± 9.5% in validation during training and 19.3 ± 9.2% in evaluation.

### 3.3. Annotation discrepancy and non-inferiority hypothesis testing

There was no significant difference between MAE_MLS-AI_ and MAE_Human_ (0.80±0.76 mm and 0.82±0.88 mm respectively; p=0.79). The disagreement of MLS-AI with the average human expert annotation of individual landmarks was 1.15 mm, while the average discrepancy of the individual human annotators amongst each other was 1.39 mm (annotator discrepancies were 1.32, 1.44, and 1.41 mm for annotators 1, 2, and 3, respectively).

The ratio of discrepancy of MLS-AI with human annotators was 0.83 (bootstrapped confidence interval at α=10^−5^ was 0.75, 0.92). Based on noninferiority hypothesis testing, the discrepancy of MLS-AI estimates was not significantly different (i.e., noninferior) from that of human annotators (p<10^−5^; Figure 3).

**Figure 3:**
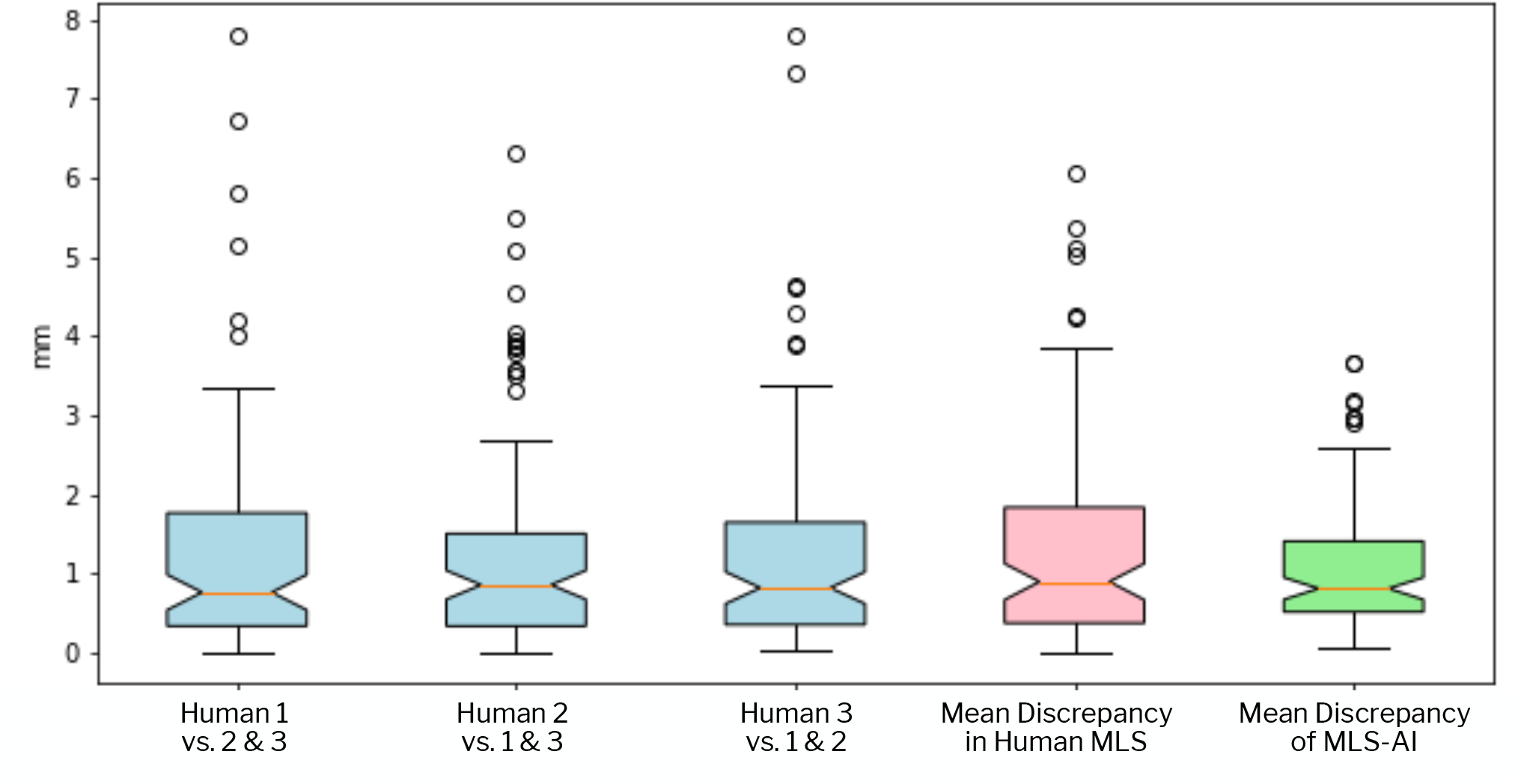
Discrepancy of MLS estimates (mm) is shown between individual human annotators and each other (cyan), between the average annotator and the individual annotators (pink), and between MLS-AI annotator (green) and the average human annotator.

### 3.4. Estimates of MLS-AI in the stroke and healthy control cohorts

The mean MLS-AI estimate was 1.37±0.14 mm in all stroke patients. The mean MLS-AI estimates for IS and HS, including IPH and SAH, were not significantly different (1.33±0.18 mm and 1.42±0.18 mm, respectively; p=0.73). See Figure 4. The mean MLS-AI estimate for the ten healthy controls was 1.01±0.41 mm, which was not significantly different from patients (p=0.07). The largest MLS-AI estimates observed in patients and in healthy subjects were 5.50 and 1.96 mm, respectively.

**Figure 4:**
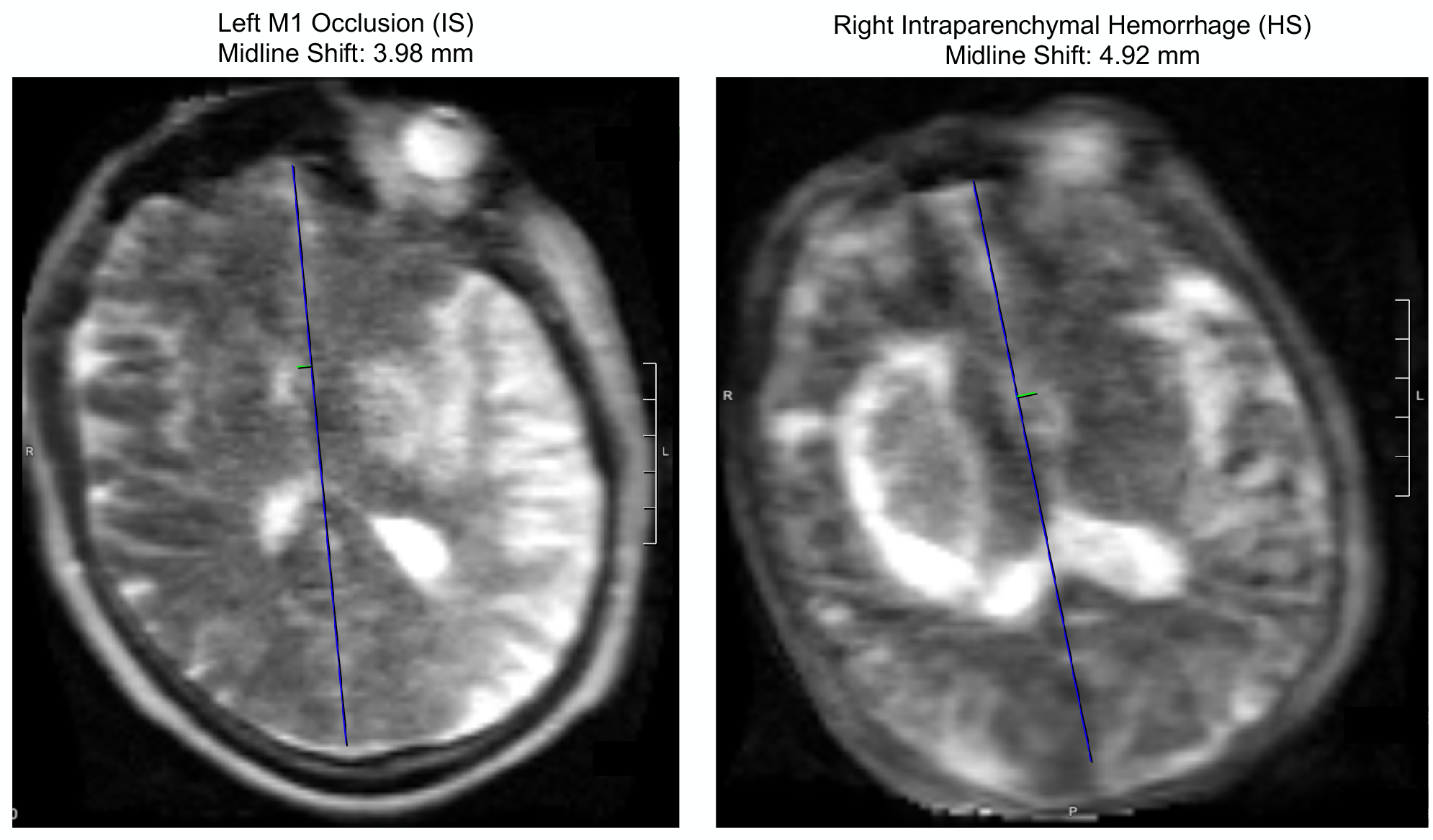
MLS-AI annotation on POC-MRI of (left) left hemisphere ischemic stroke due to occlusion in left middle cerebral artery (proximal M1 segment) and (right) right hemisphere intraparenchymal hemorrhage. Brain midline (blue) shown with midline displacement (green).

### 3.5. Relationships between MLS-AI estimates and clinical measures

MLS-AI was significantly associated with NIHSS in all patients (p<0.005) and in the IS subgroup (p=0.005) but not in the HS subgroup (p=0.13) (Figure 5b). Patient age was found to be significantly associated with NIHSS (p<0.005). In a multivariate analysis controlling for patient age, NIHSS was associated with MLS-AI in all patients and in IS (both p<0.05) but not in HS (p=0.23) (statistical summary in Supplementary Table 1).

**Figure 5:**
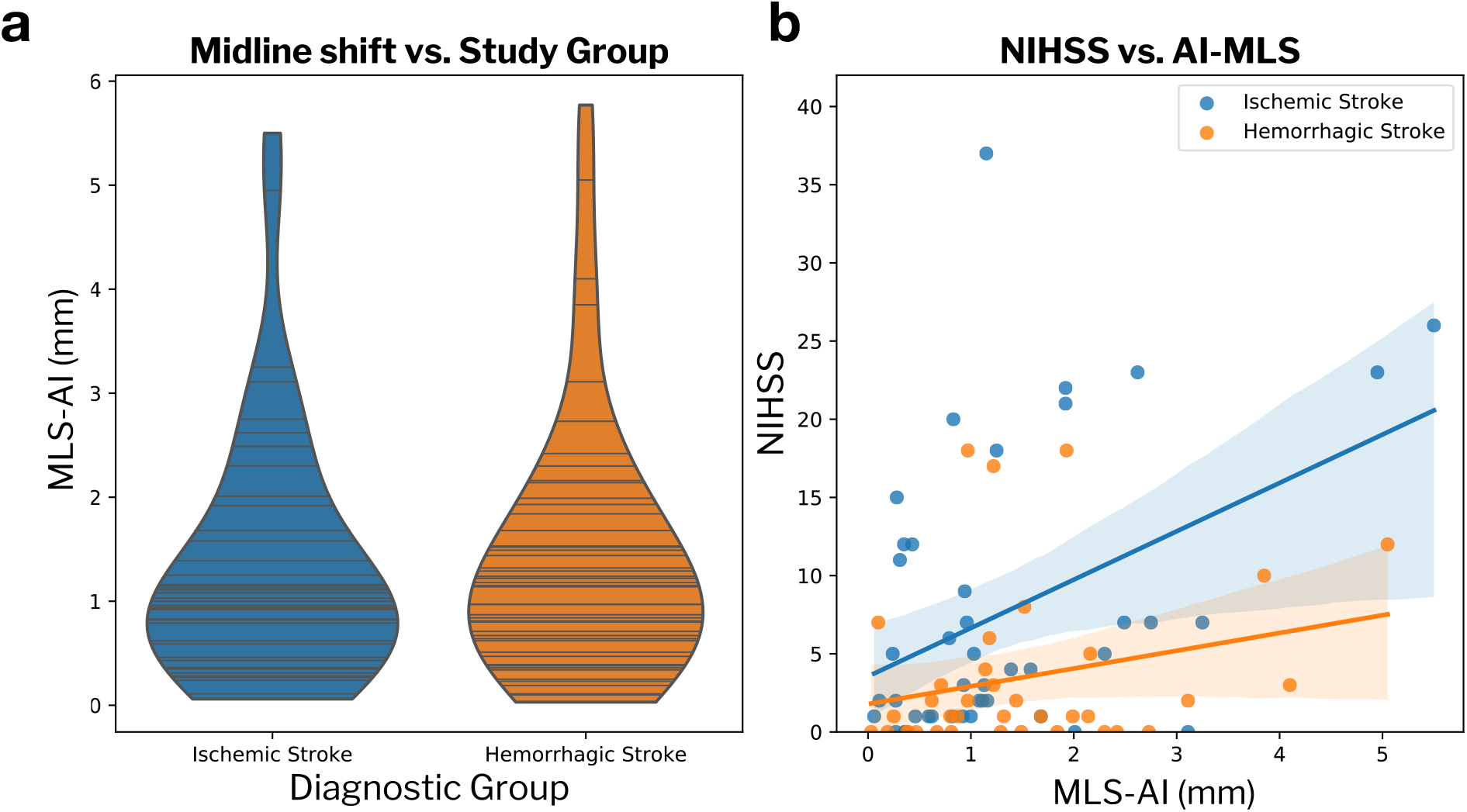
(a) Violin plots showing distributions of MLS-AI by diagnostic group: ischemic stroke (0.44 mm and 1.80 mm at first and third quartiles, respectively; n=38) and hemorrhagic stroke (0.59 mm and 1.86 mm at first and third quartiles, respectively; n=33). A two-tailed t-test indicated no significant difference in MLS-AI between patient groups (p=0.73). Horizontal lines represent individual sample points. (b) Scatterplot of MLS-AI versus stroke severity (NIHSS, 1– 42) with linear trends by study subgroup: IS (orange, β=3.093, p(β)=0.005, CI: 0.703, 3.495), HS (blue, β=0.1.137, p(β)=0.135, CI: –0.368, 2.641). The linear trend over all patients yielded parameters β=2.099, p(r)=0.004, CI: 0.703, 3.495).

In the univariate analysis, mRS was significantly correlated with MLS-AI in all patients and in the IS subgroup (both p<0.05; Figure 6) but not in HS (p=0.84). Patient age was not significantly associated with mRS, and in a multiple regression model factoring age, MLS-AI remained a significant predictor of mRS in all patients (p=0.04). For the subset of patients with follow-up data (n=26; IS:9, HS:15), a significant association of MLS-AI with 90-day mRS was observed (p<0.05: β=0.687, CI: [0.083, 1.292]).

**Figure 6.**
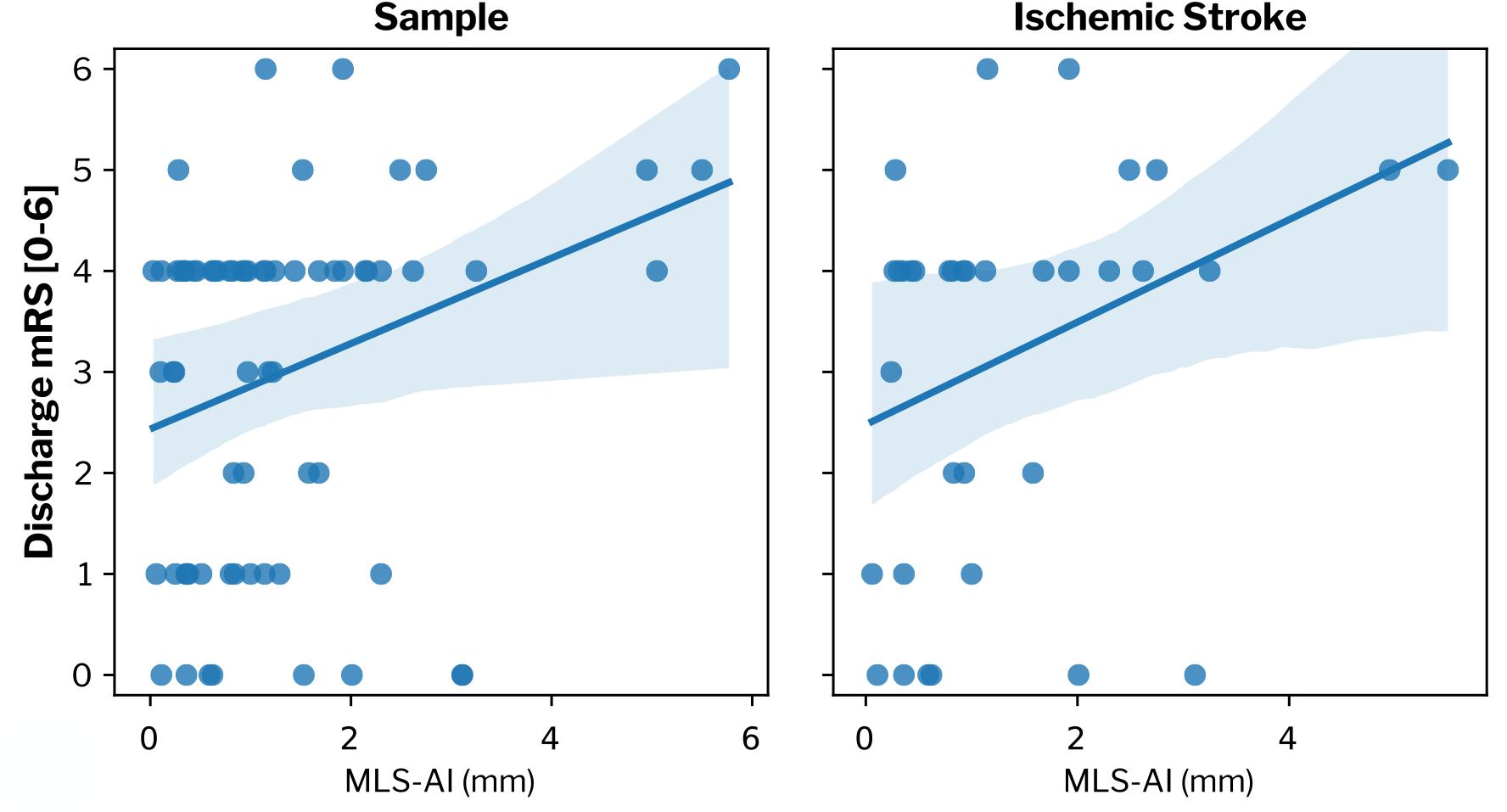
Scatterplot of morbidity at discharge according to the modified Rankin Scale (mRS; 0–6 [no disability–expiry]) versus MLS-AI with linear regression analysis in: all patients (β=0.385, p(β)=0.019, CI: 0.066, 0.704), ischemic stroke (IS) only (β=0.481, p(β)=0.040, CI: 0.023, 0.943), and hemorrhagic stroke (HS) only (β=0.062, p(β)=0.835, CI: –0.552, 0.676).

Larger MLS-AI measurements were significantly associated with worse outcome at discharge in all patients (p<0.05; OR=1.66 [CI: 1.01, 2.72]). Patient sub-groups did not show significant associations between MLS-AI and discharge outcome, with or without correcting for patient age (see Supplemental Table 1 for statistical values).

### 3.6. Qualitative threshold for MLS

Qualitatively worse disability was significantly associated with MLS-AI >1.5 mm (Mann-Whitney U-test, p<0.05). The positive predictive value (PPV) of significant disability at discharge (mRS>3; MLS-AI>1.5 mm) was 70% for the entire sample and 77% in IS. The negative predictive value (MLS-AI<1.5 mm) was marginal in both samples (51 and 46%, respectively).

## 4. Discussion

The results of our study demonstrated that MLS-AI estimates are not inferior to manual MLS measurements made by expert neuroradiologists. In addition, MLS-AI was associated with neurologic status (NIHSS) at the time of imaging and disability at discharge (mRS), before and after controlling for patient age. Furthermore, in a sub-sample of patients with follow-up data, MLS-AI was predictive of disability at 90-days post-discharge.

In practice, the Hyperfine POC-MRI transmits images to the cloud for processing, where MLS-AI models automatically evaluate new images and produce MLS-AI estimates. This approach makes MLS-AI measurement available wherever an internet connection is available, including in the developing world. The noninferiority of the MLS-AI estimates compared to expert neuroradiologist measurements suggests that clinical workflows using MLS-AI estimates could expedite evaluation of POC-MRI. Thus, the integration of AI and POC-MRI could not only save minutes to hours from image acquisition to initial interpretation, but it could also decrease healthcare costs associated with stroke, as well as increase the accessibility and utility of brain imaging.

The association of MLS-AI with outcome was observed in all patients, and specifically in IS, where swelling or edema resulting from neuronal death causes lateral shifts of midline brain structures, leading to functional disability (Adams et al., 1999; Yoo et al., 2013). Both IS and HS groups exhibited comparable distributions of MLS-AI, indicating that IS-specific associations of MLS-AI were not biased by an interaction of diagnosis and MLS-AI effect size. Importantly, no clinical outcome data was used in DL model training, suggesting that the association with outcomes were unbiased. IS accounts for approximately 87% of stroke occurrences (Ballarin and Tymianski, 2018; Beal, 2010). The stronger association of MLS-AI with IS outcomes suggested the sensitivity of this approach to brain edema. Our findings confirm that the training strategy used here rendered a model that was robust to the conditions of bedside imaging in a neurointensive care setting, suggesting a role for POC-MRI and AI in detecting stroke pathophysiology in a general stroke patient group, and in IS specifically.

Larger MLS is known to be associated with neurological deterioration and early mortality in ischemic stroke (Pullicino et al., 1997; Qureshi et al., 2009; Sandoval and Witt, 2008; Sheth et al., 2020; Wijdicks et al., 2014; Yoo et al., 2013). While the qualitative determination of significant MLS from CT and MRI has been cited as the displacement of midline structures by as much as 12 mm, shifts as small as 2 mm are associated with functional deficits (Ropper, 1986). Although the inclusion of additional predictors (e.g., gender, ethnicity, NIHSS) in models for functional outcome has been shown in stroke with larger volumes of infarction (i.e., MLS=8–22 mm), previous studies have mainly used univariate models when associating smaller MLS with functional outcomes (Battey et al., 2014; Ropper, 1986). We found that POC-MRI measures of MLS-AI greater than 1.5 mm—the voxel (i.e., volumetric pixel) size of the T2W imaging sequence used in this study—were predictive of functional outcomes. As patients in the present study were clinically stable and scanned with POC-MRI as follow up to initial treatment with appropriate intervention (i.e., thrombolytics in IS), MLS was smaller than would be observed in a typical acute stroke population. We hypothesize that MLS-AI may also be sensitive in larger strokes and when used immediately after injury.

Our study did have limitations. Motion and other artifacts corrupted 24% of the images acquired according to the clinical protocol, associated with the inclusion of lower quality images from early iterations of the POC-MRI software. Improvements in image quality and model performance are expected to improve sensitivity to MLS, and thus the prediction of outcomes. Furthermore, the predictive value for outcomes based on MLS-AI of 1.5 mm may be biased by factors such as partial volume effects, which require further study. Lastly, studying patients with more severe pathologies (i.e., larger MLS) is needed for the further validation of this approach (Battey et al., 2014). Future studies could include more patient follow-up data to demonstrate the relationship between MLS-AI and long-term outcomes and evaluate MLS as a dynamic process through serial imaging using POC-MRI.

Deep learning models used in brain imaging tasks such as segmentation and noise reduction include deep CNNs, generative adversarial networks, and autoencoders such as U-Net (Çiçek et al., 2016; Ronneberger et al., 2015). ResUNet — an evolution from U-Net and Res-Net architectures (He et al., 2016; Ronneberger et al., 2015) — leverages U-net design and has been shown to be effective in a variety of 3D image evaluation tasks (Fu et al., 2020; Wolny et al., 2020). Like all MRI, POC-MRI is susceptible to artifacts from motion and interference, where image quality can affect detection tasks. However, our results showed that ResUNet was effective in detecting anatomical landmarks in POC-MRI images while accommodating variance due to errors related to imaging in the open environment. Additionally, the MLS-AI localization accuracy metrics indicated that the MLS-AI model was not overfit and would be able to accurately identify MLS landmarks in novel data.

Limitations of two-fold cross-validation include susceptibility to selection bias and the need for data selection from the same population, in terms of subjects and data quality. These effects were controlled in this study by producing a set of different models trained with random reshuffling of data folds, and final MLS-AI estimates produced as an average of the estimates of the individual models. Using this approach, independent MLS-AI models provided unbiased MLS-AI estimates for each POC-MRI image acquired in this study.

## 5. Conclusion

Our study used an integrated approach to diagnostic brain imaging by combining portable point-of-care MRI with AI. We demonstrated the feasibility of using a POC-MRI exam to derive an automated imaging measure reflective of cerebral edema, MLS-AI, with validation against standard scores for clinical outcome, including neurologic status (NIHSS) and discharge functional outcome (mRS). The detection of MLS using POC-MRI acquired at the bedside represents a new opportunity to safely inform treatment planning throughout the progression of stroke using automated imaging methods. Further study may lead to the integration of AI and other POC-MRI-based automated imaging measures into new neurocritical care workflows to improve patient outcomes by decreasing time to treatment and increasing the accessibility of life-saving brain imaging techniques, not only in stroke, but also in other critical brain injuries.

## Data Availability

All data produced in the present study are available upon reasonable request to the authors.

## Abbreviations

AI: Artificial Intelligence
CI: Confidence Interval
DL: Deep Learning
HS: Hemorrhagic Stroke
IPH: Intraparenchymal Hemorrhage
IS: Ischemic Stroke
MLS: Midline Shift
mRS: Modified Rankin Scale
NIHSS: National Institutes of Health Stroke Scale
POC-MRI: Point-of-care MRI
PPV: Positive Predictive Value
SAH: Subarachnoid Hemorrhage
TCD: Transcranial Doppler

## Acknowledgements

This study was funded by an American Heart Association Collaborative Science Award 17CSA3355004. Additional research funding and the POC-MRI prototype was provided by Hyperfine, Inc. KNS is supported by the NIH (U24NS107136, U24NS107215, R01NR018335, R01NS110721, R03NS112859, U01NS106513, 1U01NS106513-01A1) and the American Heart Association (18TPA34170180). The authors would like to thank the volunteers for their participation and Dr. Lori Arlinghaus for assistance with manuscript preparation.

## References

Adams, H.P., Davis, P.H., Leira, E.C., Chang, K.C., Bendixen, B.H., Clarke, W.R., Woolson, R.F., Hansen, M.D., 1999. Baseline NIH Stroke Scale score strongly predicts outcome after stroke: A report of the Trial of Org 10172 in Acute Stroke Treatment (TOAST). Neurology 53, 126–131. https://doi.org/10.1212/wnl.53.1.126

Ballarin, B., Tymianski, M., 2018. Discovery and development of NA-1 for the treatment of acute ischemic stroke. Acta Pharmacol Sin 39, 661–668. https://doi.org/10.1038/aps.2018.5

Battey, T.W.K., Karki, M., Singhal, A.B., Wu, O., Sadaghiani, S., Campbell, B.C.V., Davis, S.M., Donnan, G.A., Sheth, K.N., Kimberly, W.T., 2014. Brain edema predicts outcome after non-lacunar ischemic stroke. Stroke 45, 3643–3648. https://doi.org/10.1161/STROKEAHA.114.006884

Beal, C.C., 2010. Gender and stroke symptoms: a review of the current literature. J Neurosci Nurs 42, 80–87.

Blanco, P., Abdo-Cuza, A., 2018. Transcranial Doppler ultrasound in neurocritical care. J Ultrasound 21, 1–16. https://doi.org/10.1007/s40477-018-0282-9

Çiçek, Ö., Abdulkadir, A., Lienkamp, S.S., Brox, T., Ronneberger, O., 2016. 3D U-Net: Learning Dense Volumetric Segmentation from Sparse Annotation, in: Ourselin, S., Joskowicz, L., Sabuncu, M.R., Unal, G., Wells, W. (Eds.), Medical Image Computing and Computer-Assisted Intervention – MICCAI 2016, Lecture Notes in Computer Science. Springer International Publishing, Cham, pp. 424–432. https://doi.org/10.1007/978-3-319-46723-8_49

Cooley, C.Z., McDaniel, P.C., Stockmann, J.P., Srinivas, S.A., Cauley, S.F., Śliwiak, M., Sappo, C.R., Vaughn, C.F., Guerin, B., Rosen, M.S., Lev, M.H., Wald, L.L., 2021. A portable scanner for magnetic resonance imaging of the brain. Nature Biomedical Engineering 5, 229–239. https://doi.org/10.1038/s41551-020-00641-5

Dewan, M.C., Rattani, A., Gupta, S., Baticulon, R.E., Hung, Y.-C., Punchak, M., Agrawal, A., Adeleye, A.O., Shrime, M.G., Rubiano, A.M., Rosenfeld, J.V., Park, K.B., 2018. Estimating the global incidence of traumatic brain injury. J Neurosurg 1–18. https://doi.org/10.3171/2017.10.JNS17352

Fu, F., Wei, J., Zhang, M., Yu, F., Xiao, Y., Rong, D., Shan, Y., Li, Yan, Zhao, C., Liao, F., Yang, Z., Li, Yuehua, Chen, Y., Wang, X., Lu, J., 2020. Rapid vessel segmentation and reconstruction of head and neck angiograms using 3D convolutional neural network. Nat Commun 11, 4829. https://doi.org/10.1038/s41467-020-18606-2

GBD 2015 Neurological Disorders Collaborator Group, 2017. Global, regional, and national burden of neurological disorders during 1990-2015: a systematic analysis for the Global Burden of Disease Study 2015. Lancet Neurol 16, 877–897. https://doi.org/10.1016/S1474-4422(17)30299-5

Hainc, N., Federau, C., Stieltjes, B., Blatow, M., Bink, A., Stippich, C., 2017. The Bright, Artificial Intelligence-Augmented Future of Neuroimaging Reading. Front Neurol 8, 489. https://doi.org/10.3389/fneur.2017.00489

He, K., Zhang, X., Ren, S., Sun, J., 2016. Deep Residual Learning for Image Recognition, in: 2016 IEEE Conference on Computer Vision and Pattern Recognition (CVPR). Presented at the 2016 IEEE Conference on Computer Vision and Pattern Recognition (CVPR), pp. 770–778. https://doi.org/10.1109/CVPR.2016.90

Jia, L., Wang, H., Gao, Y., Liu, H., Yu, K., 2016. High incidence of adverse events during intra-hospital transport of critically ill patients and new related risk factors: a prospective, multicenter study in China. Crit Care 20, 12. https://doi.org/10.1186/s13054-016-1183-y

LaRovere, K.L., Brett, M.S., Tasker, R.C., Strauss, K.J., Burns, J.P., Pediatric Critical Nervous System Program, 2012. Head computed tomography scanning during pediatric neurocritical care: diagnostic yield and the utility of portable studies. Neurocrit Care 16, 251–257. https://doi.org/10.1007/s12028-011-9627-3

Lau, V.I., Arntfield, R.T., 2017. Point-of-care transcranial Doppler by intensivists. Crit Ultrasound J 9. https://doi.org/10.1186/s13089-017-0077-9

LeCun, Y., Bengio, Y., Hinton, G., 2015. Deep learning. Nature 521, 436–444. https://doi.org/10.1038/nature14539

Liao, C.-C., Chen, Y.-F., Xiao, F., 2018. Brain Midline Shift Measurement and Its Automation: A Review of Techniques and Algorithms. Int J Biomed Imaging 2018. https://doi.org/10.1155/2018/4303161

Naqvi, J., Yap, K.H., Ahmad, G., Ghosh, J., 2013. Transcranial Doppler Ultrasound: A Review of the Physical Principles and Major Applications in Critical Care. International Journal of Vascular Medicine 2013, e629378. https://doi.org/10.1155/2013/629378

Ostwaldt, A.-C., Battey, T.W.K., Irvine, H.J., Campbell, B.C.V., Davis, S.M., Donnan, G.A., Kimberly, W.T., 2018. Comparative Analysis of Markers of Mass Effect after Ischemic Stroke. J Neuroimaging 28, 530–534. https://doi.org/10.1111/jon.12525

Parmentier-Decrucq, E., Poissy, J., Favory, R., Nseir, S., Onimus, T., Guerry, M.-J., Durocher, A., Mathieu, D., 2013. Adverse events during intrahospital transport of critically ill patients: incidence and risk factors. Annals of Intensive Care 3, 10. https://doi.org/10.1186/2110-5820-3-10

Peace, K., Wilensky, E.M., Frangos, S., MacMurtrie, E., Shields, E., Hujcs, M., Levine, J., Kofke, A., Yang, W., Le Roux, P.D., 2010. The use of a portable head CT scanner in the intensive care unit. J Neurosci Nurs 42, 109–116. https://doi.org/10.1097/jnn.0b013e3181ce5c5b

Pullicino, P.M., Alexandrov, A.V., Shelton, J.A., Alexandrova, N.A., Smurawska, L.T., Norris, J.W., 1997. Mass effect and death from severe acute stroke. Neurology 49, 1090–1095. https://doi.org/10.1212/wnl.49.4.1090

Quinn, T.J., Dawson, J., Walters, M.R., Lees, K.R., 2009. Reliability of the modified Rankin Scale: a systematic review. Stroke 40, 3393–3395. https://doi.org/10.1161/STROKEAHA.109.557256

Qureshi, A.I., Mendelow, A.D., Hanley, D.F., 2009. Intracerebral haemorrhage. The Lancet 373, 1632–1644. https://doi.org/10.1016/S0140-6736(09)60371-8

Rearick, T., Charvat, G.L., Rosen, M.S., Rothberg, J.M., 2017. Noise suppression methods and apparatus. US9797971B2.

Ronneberger, O., Fischer, P., Brox, T., 2015. U-Net: Convolutional Networks for Biomedical Image Segmentation, in: Navab, N., Hornegger, J., Wells, W.M., Frangi, A.F. (Eds.), Medical Image Computing and Computer-Assisted Intervention – MICCAI 2015, Lecture Notes in Computer Science. Springer International Publishing, Cham, pp. 234–241. https://doi.org/10.1007/978-3-319-24574-4_28

Ropper, A.H., 1986. Lateral displacement of the brain and level of consciousness in patients with an acute hemispheral mass. N Engl J Med 314, 953–958. https://doi.org/10.1056/NEJM198604103141504

Rumboldt, Z., Huda, W., All, J.W., 2009. Review of Portable CT with Assessment of a Dedicated Head CT Scanner. AJNR Am J Neuroradiol 30, 1630–1636. https://doi.org/10.3174/ajnr.A1603

Sandoval, K.E., Witt, K.A., 2008. Blood-brain barrier tight junction permeability and ischemic stroke. Neurobiol Dis 32, 200–219. https://doi.org/10.1016/j.nbd.2008.08.005

Seabold, S., Perktold, J., 2010. Statsmodels: Econometric and Statistical Modeling with Python. Proceedings of the 9th Python in Science Conference 92–96. https://doi.org/10.25080/Majora-92bf1922-011

Serag, A., Ion-Margineanu, A., Qureshi, H., McMillan, R., Saint Martin, M.-J., Diamond, J., O’Reilly, P., Hamilton, P., 2019. Translational AI and Deep Learning in Diagnostic Pathology. Front Med (Lausanne) 6, 185. https://doi.org/10.3389/fmed.2019.00185

Sheth, K.N., Mazurek, M.H., Yuen, M.M., Cahn, B.A., Shah, J.T., Ward, A., Kim, J.A., Gilmore, E.J., Falcone, G.J., Petersen, N., Gobeske, K.T., Kaddouh, F., Hwang, D.Y., Schindler, J., Sansing, L., Matouk, C., Rothberg, J., Sze, G., Siner, J., Rosen, M.S., Spudich, S., Kimberly, W.T., 2020. Assessment of Brain Injury Using Portable, Low-Field Magnetic Resonance Imaging at the Bedside of Critically Ill Patients. JAMA Neurol. https://doi.org/10.1001/jamaneurol.2020.3263

Smith, I., Fleming, S., Cernaianu, A., 1990. Mishaps during transport from the intensive care unit. Crit Care Med 18, 278–281. https://doi.org/10.1097/00003246-199003000-00006

Sulter, G., Steen, C., De Keyser, J., 1999. Use of the Barthel index and modified Rankin scale in acute stroke trials. Stroke 30, 1538–1541. https://doi.org/10.1161/01.str.30.8.1538

Taghanaki, S.A., Abhishek, K., Cohen, J.P., Cohen-Adad, J., Hamarneh, G., 2021. Deep semantic segmentation of natural and medical images: a review. Artif Intell Rev 54, 137– 178. https://doi.org/10.1007/s10462-020-09854-1

Turpin, J., Unadkat, P., Thomas, J., Kleiner, N., Khazanehdari, S., Wanchoo, S., Samuel, K., Moclair, B.O., Black, K., Dehdashti, A.R., Narayan, R.K., Temes, R., Schulder, M., 2020. Portable Magnetic Resonance Imaging for ICU Patients. Crit Care Explor 2, e0306. https://doi.org/10.1097/CCE.0000000000000306

Van Essen, D.C., Smith, S.M., Barch, D.M., Behrens, T.E.J., Yacoub, E., Ugurbil, K., WU-Minn HCP Consortium, 2013. The WU-Minn Human Connectome Project: an overview. Neuroimage 80, 62–79. https://doi.org/10.1016/j.neuroimage.2013.05.041

Vieira, S., Pinaya, W.H.L., Mechelli, A., 2017. Using deep learning to investigate the neuroimaging correlates of psychiatric and neurological disorders: Methods and applications. Neurosci Biobehav Rev 74, 58–75. https://doi.org/10.1016/j.neubiorev.2017.01.002

Walker, E., Nowacki, A.S., 2011. Understanding Equivalence and Noninferiority Testing. J Gen Intern Med 26, 192–196. https://doi.org/10.1007/s11606-010-1513-8

Wijdicks, E.F.M., Sheth, K.N., Carter, B.S., Greer, D.M., Kasner, S.E., Kimberly, W.T., Schwab, S., Smith, E.E., Tamargo, R.J., Wintermark, M., American Heart Association Stroke Council, 2014. Recommendations for the management of cerebral and cerebellar infarction with swelling: a statement for healthcare professionals from the American Heart Association/American Stroke Association. Stroke 45, 1222–1238. https://doi.org/10.1161/01.str.0000441965.15164.d6

Wolny, A., Cerrone, L., Vijayan, A., Tofanelli, R., Barro, A.V., Louveaux, M., Wenzl, C., Strauss, S., Wilson-Sánchez, D., Lymbouridou, R., Steigleder, S.S., Pape, C., Bailoni, A., Duran-Nebreda, S., Bassel, G.W., Lohmann, J.U., Tsiantis, M., Hamprecht, F.A., Schneitz, K., Maizel, A., Kreshuk, A., 2020. Accurate and versatile 3D segmentation of plant tissues at cellular resolution. Elife 9, e57613. https://doi.org/10.7554/eLife.57613

Yoo, A.J., Sheth, K.N., Kimberly, W.T., Chaudhry, Z.A., Elm, J.J., Jacobson, S., Davis, S.M., Donnan, G.A., Albers, G.W., Stern, B.J., González, R.G., 2013. Validating imaging biomarkers of cerebral edema in patients with severe ischemic stroke. J Stroke Cerebrovasc Dis 22, 742–749. https://doi.org/10.1016/j.jstrokecerebrovasdis.2012.01.002

Yushkevich, P.A., Yang Gao null,, Gerig, G., 2016. ITK-SNAP: An interactive tool for semi-automatic segmentation of multi-modality biomedical images. Annu Int Conf IEEE Eng Med Biol Soc 2016, 3342–3345. https://doi.org/10.1109/EMBC.2016.7591443

