## Supplemental Materials for "Point-of-Care MRI with Artificial Intelligence to Measure Midline Shift in Acute Stroke Follow-Up"

**Supplemental Methods**

*Training data augmentation*

Training images were used as a basis for data augmentation. Affine transformations were applied with the following parameters to augment image content:

- scale (0.9, 1.1)
- rotation (degrees): (–5, 5)
- translation (in pixels): (–3,3)

A synthetic shift was then applied to the image using an affine transform in order to increase the variability of midline shifts to train the model for robustness. For training data adapted from high-field MRI, image quality was adapted by first transforming the image volume to k-space in order to adapt POC-MRI spatial frequency encoding. Ten percent of the k-space data was randomly dropped, creating aliasing. The data were then transformed back to the image domain and Gaussian noise was added such that the signal-to-noise ratio (SNR) of the image was set to be in range (9, 13) dB, matching POC-MRI SNR.

The anatomical landmarks of the falx cerebri and septum pellucidum were represented as 3D image coordinates. The same rescaling, affine transformations, and deformation transformations were applied to the coordinate points to maintain the correspondence between augmented data and the target coordinate points.

The coordinate points were then transformed to heatmap representations using a full-width-half-max filter. These resulting images were the target for the DL training as a tensor of shape (112, 112, 96, 3), where each landmark was assigned its own channel and the last dimension reflected the number of landmarks.

*MLS-AI Network Design*

MLS-AI is an end-to-end fully convolutional neural network (CNN) based on the ResUNet architecture ^1^ (Supplemental Figure 1). The network architecture used in the present study is a 3D autoencoder based on both U-net^1^ and residual network^2^ architectures (Figure 1b and c). The input to the network was a tensor of shape (112, 112, 96, 1), representing the normalized image volume. These inputs were POC-MRI image data converted from digital imaging and communications in medicine (DICOM) format, uploaded from POC-MRI. Image intensity was normalized by clipping outside of the 1^st^ and top 99^th^ percentiles and linearly normalizing values between 0 and 1. Image dimensions were then standardized to a resolution of (112, 112, 96). The output of the network was the tensor of shape (112, 112, 96, 3), which were AI generated heatmap representations of the MLS-AI annotations.

Each convolutional layer in Supplemental Figure 1 is represented as “Conv3D(k, s, n_ch_)”, which indicates a 3D convolution with kernel size *k*, stride *s*, and number of features n_ch_. Bias was used for all convolutional layers and SeLU^3^ for the nonlinear activation. The network builds a convolutional representation with multiple channels using residual blocks at multiple scales. At each scale, residual blocks were applied. When downsampled, the image dimension was halved, but the number of features were doubled. After 5 cascading layers of downsamplings, the network features were upsampled to the original size. The output at each scale was summed with the upsampled feature from the lower scale. The maximum layer depth of this network was 12. Finally, a 1 x 1 x 1 convolution was performed to assign the number of channels to match the number of target classes. The network was implemented in TensorFlow (version 1.4) ^4^.

The model was optimized using the Adam optimizer according to the following loss function, representing the mean squared error (MSE) between ground truth and predicted probability maps.

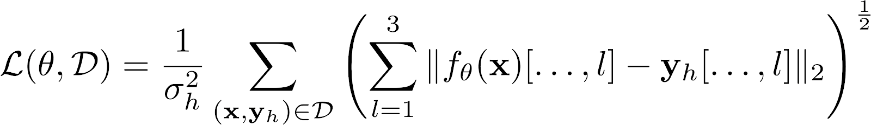

Where *l* was index for landmark, f_θ_(x) is the predicted heatmap for landmark *l*, y_h_ and is ground truth for landmark *l*. Model optimization was performed as follows: L2-regularization of the network weight was used with scale 3×10^–5^ and Polyak decay with factor 0.99.

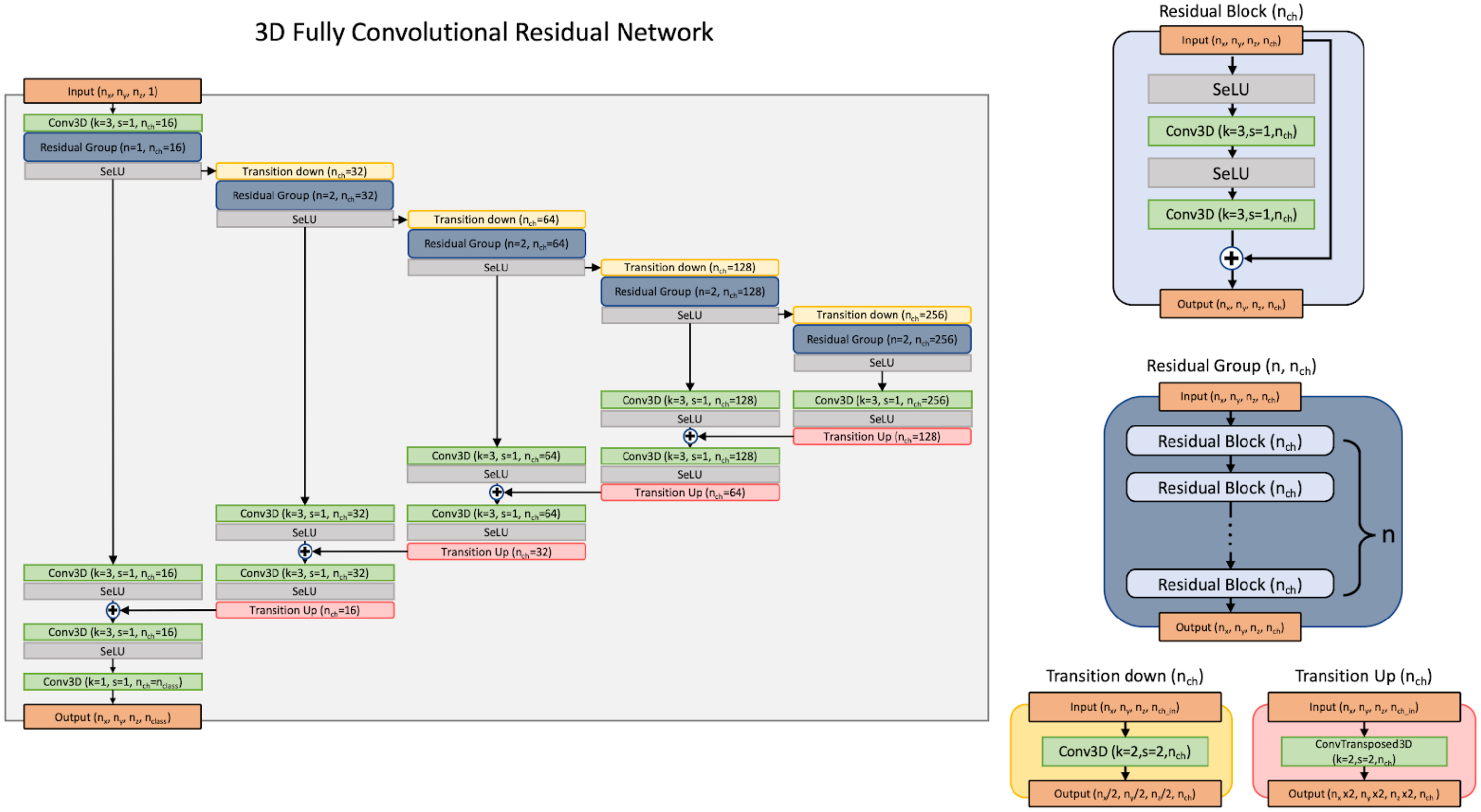

**Supplemental Figure 1:** (a) The U-net network architecture of the BrainInsight deep-learning model used in the present study, 3D ResUNet (see the main body for more details), is effective in image processing tasks owing to the preservation of spatial relationships of weights throughout the cascading layers. (b) The Res-Net components of the model architecture mitigate information loss from downsampling operations within encoding-decoding blocks of the same size.

*MLS-AI model evaluation and estimation*

The evaluation of POC-MRI volumes using trained models produced probability maps for each of the target anatomical landmarks. MLS-AI was then estimated as a vector with the length of perpendicular distance (mm) between the peak probability for the septum pellucidum to the projection brain midline, mimicking the approach to human measurement, producing a 3D vector between the connection points of anterior and posterior falx cerebri.

The input tensor was subsequently fed to the 10 trained models in the ensemble. Each model generated the output of shape (112, 112, 96, 3), which were the predicted heatmaps for each landmark. Each output landmark coordinate was calculated by taking the *argmax* of the heatmap, and was subsequently transformed back to the original coordinate system in NifTI image format using the inverse affine transformation.

The evaluation of a model group representing a data fold was done by ensembling based on simple averaging of the predicted coordinate points with outlier rejection via the angle-based outlier detection from PyOD ^5^. The final output was given by the average coordinate points of the 6 most inliers. The output of the ensemble was the 3D coordinate points of the three landmarks.

The final MLS-AI measurement of the algorithm was calculated from the ensemble output as the shortest distance between the septum pellucidum and the midline that passed through anterior falx and posterior falx. Sanity checks were performed to ensure that the output MLS-AI was valid. The slope of the line passing through the anterior falx and posterior falx was calculated and the output rejected if the slope deviated more than 10 degrees from the anterior-posterior axis. The location of the septum pellucidum was also calculated and the output rejected if the landmark was 50–150 mm outside of the field of view of the superior-inferior axis. Finally, the output was rejected if an MLS measurement was greater than 40 mm.

*Assessment of discrepancy and non-inferiority between MLS-AI and annotators*

When assessing the accuracy of a measurement made by a human or AI annotator, there is typically a ground truth quantity available. In such instances, discrepancy between ground truth and any estimate can be obtained and then averaged over a dataset to obtain an overall assessment of discrepancy ^6^. While many error metrics exist for comparing an estimate with ground truth, we use the mean absolute error (MAE) for our measure of discrepancy of linear measurements.

As different annotations produce different measurement values given the same MRI brain image, obtaining “true” ground truth is intractable since the observed inter-annotator variation is evidence that even trained clinicians are imperfectly measuring the underlying measurements. Consequently, measuring both model and annotator discrepancy against ground truth is not directly computable. Instead, it must be approximated. The following sections outline how such an approximation is made in order to compute the discrepancy of both annotators and the model.

As different human annotators produce different measurement values given the same MRI brain image, comparison to “true” ground truth is intractable since the observed inter-annotator variation is evidence that even trained clinicians are imperfectly measuring the underlying measurements. In order to evaluate an individual annotator’s discrepancy on a given anatomical measurement (i.e., landmark), an annotator’s estimate (human or MLS-AI) was compared to the approximate ground truth estimate established by the estimates provided by the *other* annotators for a given anatomical measurement (i.e., landmark).

Formally, let
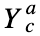
 represent the measurement provided by annotator *a* for a brain image *c* and 
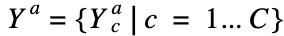
 represent the set of measurements by annotator *a* over every brain image. For a given annotator, we compute the MAE of linear measurements for our measure of discrepancy via:

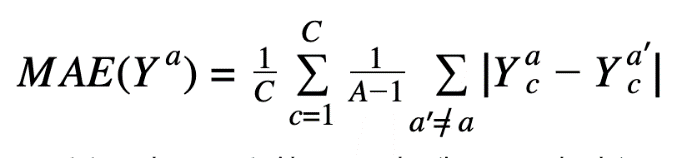

Note that the MAE for annotator *a* is computed by averaging the mean absolute error between each of their measurements and every other annotator for a given brain image. The greater the MAE score, the greater the discrepancy.

In order to compute an annotator’s discrepancy on a set of *n* 3D landmarks, for a given annotator, we compute his or her MSE of *n*x3 dimensional vector for our measure of discrepancy as follows:

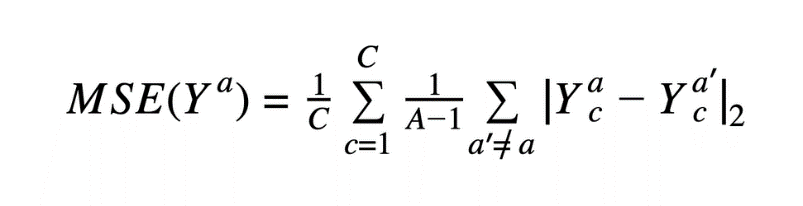

where 
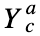
 represents the concatenation of all 3D landmarks provided by annotator *a* for a brain image *c.*

*Establishing non-inferiority*

After assessing annotator and model discrepancy, the model was validated in terms of non-inferiority^7^ to that of human experts in its ability to estimate a linear measurement. Note, the test we outline here does not preclude a model from possibly being superior to annotators. However, our methodology is geared towards validating that our algorithm is just as good as skilled human radiologists.

Non-inferiority is established when we can conclude with a high degree of certainty that our model’s discrepancy in estimating linear measurements of 3D landmarks is not significantly worse than those of the average annotator. Here, when we say that the model’s discrepancy is not significantly worse than that of the average annotator, we mean that the difference in model discrepancy and the average annotator discrepancy, as a fraction of average annotator discrepancy, is upper bounded by a clinically acceptable threshold, denoted as δ. In other words, we establish non-inferiority by showing that the model’s discrepancy is significantly less than (1 + δ) x average annotator discrepancy.

Concretely, we can establish non-inferiority at a 0.05 significance level by showing that the upper limit on a 90% confidence interval is less than or equal to δ. We use a 90% CI rather than 95% since the test is one-sided. As long as we ensure that the null distribution of Δh has no more than 5% mass greater than δ (which is what the upper limit on a 90% CI will show), we can reject at ɑ=0.05 significance.

Having a high degree of non-inferiority implies that it is statistically unlikely that a model’s low discrepancy is an artifact of the data sample (statistical likelihood). In other words, the p-value that results from testing the hypothesis that our model’s low discrepancy was the result of chance, and should be low (p<0.05)^7^.

Formally, the hypothesis test establishes non-inferiority as follows:

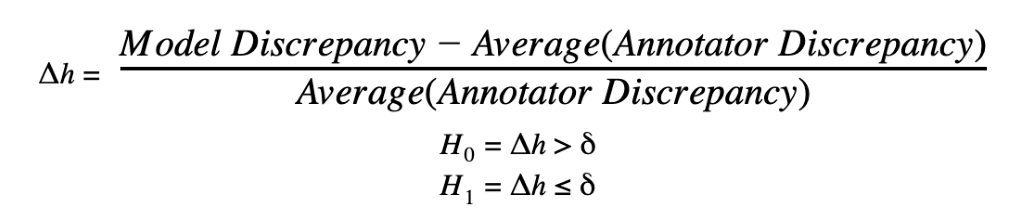

We can reject the null hypothesis at significance level α if the upper limit on the (1–2𝛼) x 100% CI on Δh is less than or equal to δ. In order to compute a CI on Δh, we bootstrap B = 10,000 trials by sampling *with replacement* C samples from the set of samples. A (1–2α) x 100% CI is found by taking the α x 100% and (1–α) x 100% quantiles from the set of B test-statistics. If the (1–α) x 100% quantile is < δ, we can reject the null hypothesis at p < α significance.

Intuitively, δ should be significantly less than 1.0 while still providing a region around the average observed annotator efficacy for which performance is considered non-inferior.

**Supplemental Results**

|  | **Neurologic Status**  **(NIHSS: 1–42)** | | | | **Discharge Disability**  **(mRS: 1–6)** | | | | **Outcome**  **(mRS>3)** | | | |
| --- | --- | --- | --- | --- | --- | --- | --- | --- | --- | --- | --- | --- |
|  | *Reg. Coeff.* | *[0.025* | *0.975]* | p-value | *Reg. Coeff.* | *[0.025* | *0.975]* | p-value | *Reg. Coeff.* | *[0.025* | *0.975]* | p-value |
| **IS+HS** | 2.099 | 0.703 | 3.495 | 0.004** | 0.385 | 0.066 | 0.704 | 0.019* | 0.5054 | 0.007 | 1.004 | 0.047* |
| **IS** | 3.093 | 0.973 | 5.213 | 0.005* | 0.483 | 0.023 | 0.943 | 0.040* | 0.636 | -0.137 | 1.408 | 0.107 |
| **HS** | 1.137 | -0.368 | 2.641 | 0.134 | 0.0622 | -0.552 | 0.676 | 0.835 | 0.39 | -0.456 | 1.236 | 0.366 |
| **Age** | 0.199 | 0.083 | 0.315 | 0.001** | -0.015 | -0.069 | 0.039 | 0.566 | 0.0214 | -0.012 | 0.055 | 0.209 |
|  | *Multivariate (Controlling for Age)* | | | | | | | | | | | |
| **IS+HS** | 1.96 | 0.324 | 3.606 | 0.020* | 0.332 | 0.012 | 0.651 | 0.042* | 0.4761 | -0.028 | 0.98 | 0.064 |
| **IS** | 2.4092 | 0.262 | 4.719 | 0.030* | 0.3641 | -0.101 | 0.829 | 0.121 | 0.613 | -0.005 | 0.09 | 0.154 |
| **HS** | 1.1168 | -0.812 | 3.045 | 0.24 | 0.1122 | -0.534 | 0.758 | 0.72 | 0.6215 | -0.335 | 1.578 | 0.203 |

**Supplemental Table 1:** Regression analysis demonstrating the association between stroke outcomes and midline shift analyzed by artificial intelligence (MLS-AI). Cohorts for analysis include the full sample of patients with diagnostic data (IS+HS), and IS and HS sub-groups (n=42 and n=33, respectively). Asterisks (*) indicate significant associations of patient outcomes with MLS-AI at the p<0.05 level, and (**) at the p<0.005 level. Associations were determined using ordinary least squares (OLS) regression model: $Y=\boldsymbol{\beta*X}+\boldsymbol{\varepsilon}$. IS: ischemic stroke, HS: hemorrhagic stroke, NIHSS: NIH stroke scale, mRS: modified Rankin scale for neurologic disability.

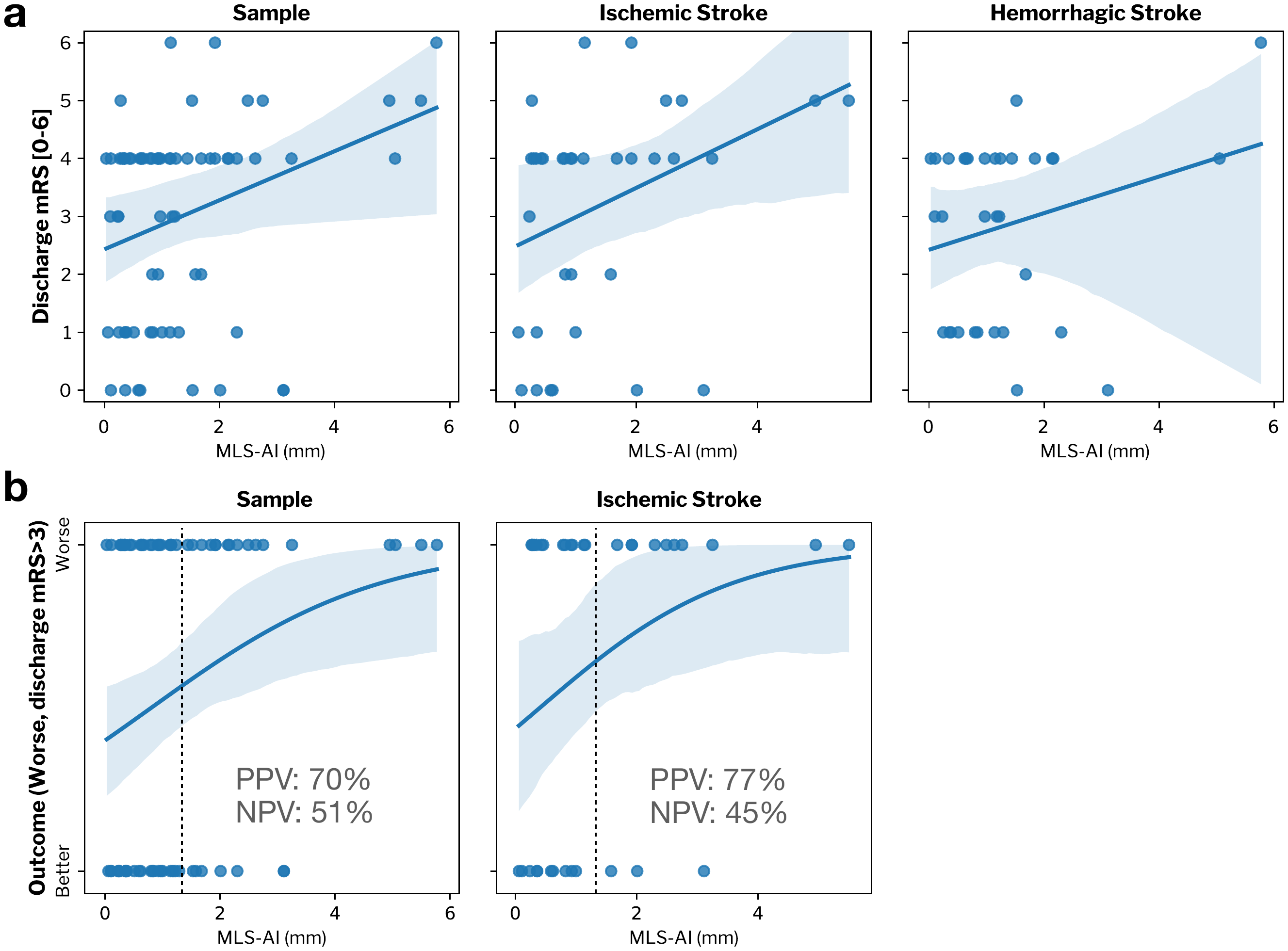

**Supplemental Figure 2:** Inpatient MLS-AI versus morbidity at subsequent discharge. (a) Scatterplot of morbidity at discharge according to the modified Rankin Scale (mRS; 0–6 [no disability–expiry]) versus MLS-AI with linear regression analysis in: all patients (β=0.385, p(β)=0.019, CI: 0.066, 0.704), ischemic stroke (IS) only (β=0.481, p(β)=0.040, CI: 0.023, 0.943), and hemorrhagic stroke (HS) only (β=0.062, p(β)=0.835, CI: –0.552, 0.676). Confidence intervals for linear trends (95%) are shown as blue overlap from leave-one-out cross-validation. (b) Scatterplot of outcome (mRS>3 or significant disability) versus MLS-AI with logistic regression analysis. Confidence intervals (95%) are shown as blue overlays for logistic regression for all patients (log(β)=0.505, p(log[β])=0.047, CI:0.007, 1.004). Positive predictive value (PPV) and negative predictive value (NPV) are shown.

**References**

1. Ronneberger, O., Fischer, P. & Brox, T. U-net: Convolutional networks for biomedical image segmentation. in *International Conference on Medical image computing and computer-assisted intervention* 234–241 (Springer, 2015).
2. Çiçek, Ö., Abdulkadir, A., Lienkamp, S. S., Brox, T. & Ronneberger, O. 3D U-net: learning dense volumetric segmentation from sparse annotation. *arXiv:1606.06650 [cs]* (2016).
3. Klambauer, G., Unterthiner, T., Mayr, A. & Hochreiter, S. Self-normalizing neural networks. *Advances in neural information processing systems* **30**, 971–980 (2017).
4. Abadi, M. *et al.* Tensorflow: a system for large-scale machine learning. in 265–283 (2016).
5. Zhao, Y., Nasrullah, Z. & Li, Z. Pyod: A python toolbox for scalable outlier detection. *arXiv preprint arXiv:1901.01588* (2019).
6. Lovchinsky, I. *et al.* Discrepancy ratio: evaluating model perfor- mance when even experts disagree on the. 19 (2020).
7. Walker, E. & Nowacki, A. S. Understanding equivalence and noninferiority testing. *J Gen Intern Med* **26**, 192–196 (2011).
